# The impact of motor, non-motor, and social aspects on the sexual health of men living with Parkinson’s disease

**DOI:** 10.1101/2023.09.21.23295906

**Authors:** Bruno Rafael Antunes Souza, Kátia Cirilo Costa Nóbrega, Bruno Eron de Almeida da Silva, Raissa Amorim Gonçalves, Thalyta Silva Martins, Geovanna Ferreira Santos, Andre Helene Frazão, Antonio Carlos Roque, Isaíra Almeida Pereira da Silva Nascimento, Maria Elisa Pimentel Piemonte

## Abstract

**Background:** Sexual health (SH) is influenced by several biological, mental, and social factors which may be negatively impacted by PD. Despite its prevalence and relevance for quality of life, the factors that affect the SH in men with Parkinson’s disease (MwPD) are still poorly understood, and research in this area is scarce.

**Aim:** To investigate the impact of motor, non-motor, and social aspects on the SH in MwPD.

**Method:** We conducted a cross-sectional study of 80 men (mean age 53.55±10.8) in stages 1-3 of disease progression according to Hoehn and Yahr classification (H&Y), who reported to have sexual activity. The following data were collected for each person (used tests/scales indicated within parentheses and defined in glossary): (1) demographic information and global cognitive capacity (T-MoCA); (2) non-motor aspects of daily life experiences (MDS-UPDRS, part I); (3) motor aspects of daily life experiences (MDS-UPDRS, part II); (4) fatigue (FSS); (5) self-esteem (RSES); (6) sleep disorder (PDSS); (7) couple relationship quality (DAS); (8) depressive signals (BDI); (8) short-term sexual health by International Index of Erectile Function (IIFE); and (9) long-term sexual health by Sexual Quotient-Male (SQ-M).

**Results:** Our results suggest that the quality of marital relationships predicts short-term SH, motor disability level predicts erectile dysfunction, and depression predicts long-term SH in MwPD. Age, disease onset, disease duration, Levodopa dosage, and non-motor symptoms, excluding depression, were not correlated with SH.

**Conclusion:** Our findings confirm that multidimensional factors can affect the SH of MwPD and emphasize that only a multi-professional team can offer proper care to improve SH in MwPD.

## Introduction

Parkinson’s disease (PD) is an idiopathic, neurodegenerative, multisystemic, progressive, and irreversible disorder with a large spectrum of motor and non-motor signs and symptoms [1]. Its incidence increases with aging and rises sharply after 65 [2]. According to the Global Burden of Disease study, PD was the only neurological disorder with increasing age-standardized rates of deaths, prevalence, and disability-adjusted life-years between 1990 and 2015 [3].

According to World Health Organization, Sexual health (SH) is “a state of physical, emotional, mental and social well-being in relation to sexuality” [4]. SH involves many factors, including organic causes, environmental influences, and psychological factors. It can be a complex phenomenon, making it difficult to study and understand fully [5].

Among the several non-motor functions impaired by PD, sexual function [6,7], and sexual self-esteem [8], key aspects of SH are negatively affected. Sexual dysfunction (SD) significantly reduces the quality of life of people with PD and increases the medical, social, and economic burden for patients, sex partners, and their caregivers [9]. However, they receive little attention from health professionals and are underreported [8,10]. Raciti et al. (2020) showed that about 57% of people stated that the onset of PD affected their sex life, mainly due to a decrease in sexual desire and the frequency of sexual intercourse [11]. A cohort study showed that people with PD showed a progressive reduction in sexual activity from 56.3% to 50.8% over 2 years of follow-up [12]. An important retrospective study over 14 years on 32,616 men with erectile dysfunction (ED) aged between 40 and 70 years showed that there was a 3.8 times higher incidence of developing PD in later years, suggesting that ED may be a prodromal symptom of PD [13]. Furthermore, the presence of SD before the PD diagnosis predicted post-diagnosis SD in men [14].

Studies confirm that SD is more prevalent in people with PD than healthy individuals [15,16]. Vela-Desojo et al. (2020) showed that only 35.2% of people with early onset of PD (EOPD) against 81.2% of healthy people reported sexual satisfaction [17]. Regarding age, studies show different results: some show that SD is worse in older people with PD (17,18,19,20,21] while others show that younger people report more DS [22] or that DS is age-independent [20]. Pedro et al. (2020) still found that 83.3% of men with PD (MwPD) treated with Deep Brain Stimulation of the subthalamic nucleus have ED, with age being an independent predictive factor for its onset [20].

Regarding disease progression and disease severity, some studies showed that SD increases with disease progression [11,23,24] and disease severity [24,25,26,27], while others showed no correlation between disease progression [18,22,28,29] or disease severity [11,19,28,30,31]. In MwPD, disease severity was associated with libido loss [26].

By gender, previous studies show a prevalence ranging between 42% and 79% in men and between 36–87% in women [6,26,31,32]. MwPD, report more SD than women [33,34] and more dissatisfaction than controls [6]. Furthermore, it has been found that hypersexuality, which is characterized by heightened sexual desire and repetitive actions aimed at achieving sexual satisfaction outside of socially or personally acceptable limits, is more common in MwPD with a prevalence of 5.2% compared to women with PD (0.5%) [35].

Frequency of intercourse, sexual arousal, subjective abnormal sexual fantasies, or sexual satisfaction deteriorated in both genders, but especially in MwPD [21]. The main SD observed in MwPD were (1) hypoactive sexual desire disorder reported by 83% of participants [32]; (2) Erectile dysfunction (ED) reported from 42.6% to 79% of MwPD according to the study [6,14,26,32,36]; (3) difficulty reaching orgasm reported by 39,5% [6], premature ejaculation reported by from 40.6% to 79% of Mw PD [6,10,37]; (5) anejaculation reported by 87% of MwPD [32] and (6) Hypersexuality reported by 5.2% of MwPD [35].

Several alterations in sensory, motor, mental, and autonomic functions associated with PD and social aspects can directly or indirectly affect the SH [12,38].

People with PD can experience motor alterations that may negatively impact their SH. These alterations may include rigidity, tremor, immobility in bed, and incoordination in fine movements of the hands and fingers. These challenges can make it difficult to experience pleasure and sexual arousal and may prevent the person with PD from being able to participate fully in sexual activity. Additionally, facial hypomimia can make it appear as though the person lacks affection and desire, which can further complicate matters [6]. In fact, people with PD showed higher levels of motor symptoms severity and motor disability than those without SD [11]. Reduced motor disability level in MwPD is associated with higher SH [8,12,37] and sexual satisfaction [39]. Interestingly, motor symptom severity predicted the higher quality of SH of the partner but not of the MwPD [7]. In MwPD, motor symptom severity was associated with ED [18]. Postural instability is a remarkable motor symptom of the intermediate PD stage, predicted SD [37].

Among the non-motor alterations, depression, and anxiety have been identified as the factor most correlated with SD (11,17,18,37,39]. For MwPD, lower depression is associated with higher sexual activity [12], and depression, age, and presence of a partner [36] or depression, and subjective scoring of the illness [39] were able to predict their SH. Furthermore, the main predictor factor for libido loss and decreased sexual desire in people with PD [26]. Özcan et al. (2015) found an association between anxiety, but not depression, and SH [18].

Few studies have investigated the impact of cognitive changes on the SH of people with PD. Hand et al. (2010) found no correlation between cognitive status and PD [22]. Picillo et al. (2019) found similar results: the SH of men and women with PD was not correlated with cognitive ability [12]. In contrast, Kummer, Cardoso, and Teixeira (2009) found an association between cognitive decline and libido loss in MwPD [26]. Urinary dysfunction has also been associated with DS complaints and decreased sexual satisfaction in people with Parkinson’s [17]. Autonomic dysfunction was associated with loss of libido in people with PD, and fatigue severity was associated with libido loss in MwPD [26].

Among the social factors, the quality of the marital relationship has been bidirectionally related to SH [34]. Brown et al. (1990) showed that couples where one of the partners had PD who reported a better marital relationship were more satisfied with their sex life and vice versa [8]. However, satisfaction with the marital relationship varied between genders: dissatisfaction with the marital relationship and the sex life of both partners was greater when the person with PD was a man. Other studies found similar results: although MwPD rejected sex less often and their sexual desire was higher than women, they reported more dissatisfaction with their sex life than women [6,7].

SH is a complex phenomenon in which, besides organic causes, environmental and psychological factors may also be involved. SD is a prevalent and disturbing non-motor symptom of PD, mainly for men. This study aimed to investigate motor, non-motor, and social aspects of the sexual health of MwPD.

## Materials and Methods

### Study Design and Participants

A cross-sectional study included 80 MwPD. The eligibility criteria were (a) man; (b) confirmed diagnosis of idiopathic PD offered by a neurologist according to the UK Parkinson’s Disease Society Brain Bank diagnostic criteria [40], age above 21 years; (c) reporting active sexual function in the last six months; and (d) have access to telephone or internet and agree to participate in the study. The non-eligibility criteria were (a) the presence of neurological disorders other than PD; and (b) the presence of dementia, speech, and hearing disorders that could impair the remote interview.

### Recruitment

Participants were recruited by phone contact. Initial eligibility was identified through calls. Subsequently, information about the study procedures was passed on, and they were invited to consent to participate.

This study was approved by the proper Ethics Committee and conducted in accordance with the Helsinki Declaration.

### Study Procedures

Interviews were conducted using a structured form and validated instruments. Participants were asked to indicate the best day and time for the remote interview.

The interviews were carried out by the same interviewer in two separate sessions, each lasting around 30-35 minutes. This was done to ensure the participants did not get tired during the process. The gap between the two interviews was kept less than 7 days.

The study flow is demonstrated in Figure 1.

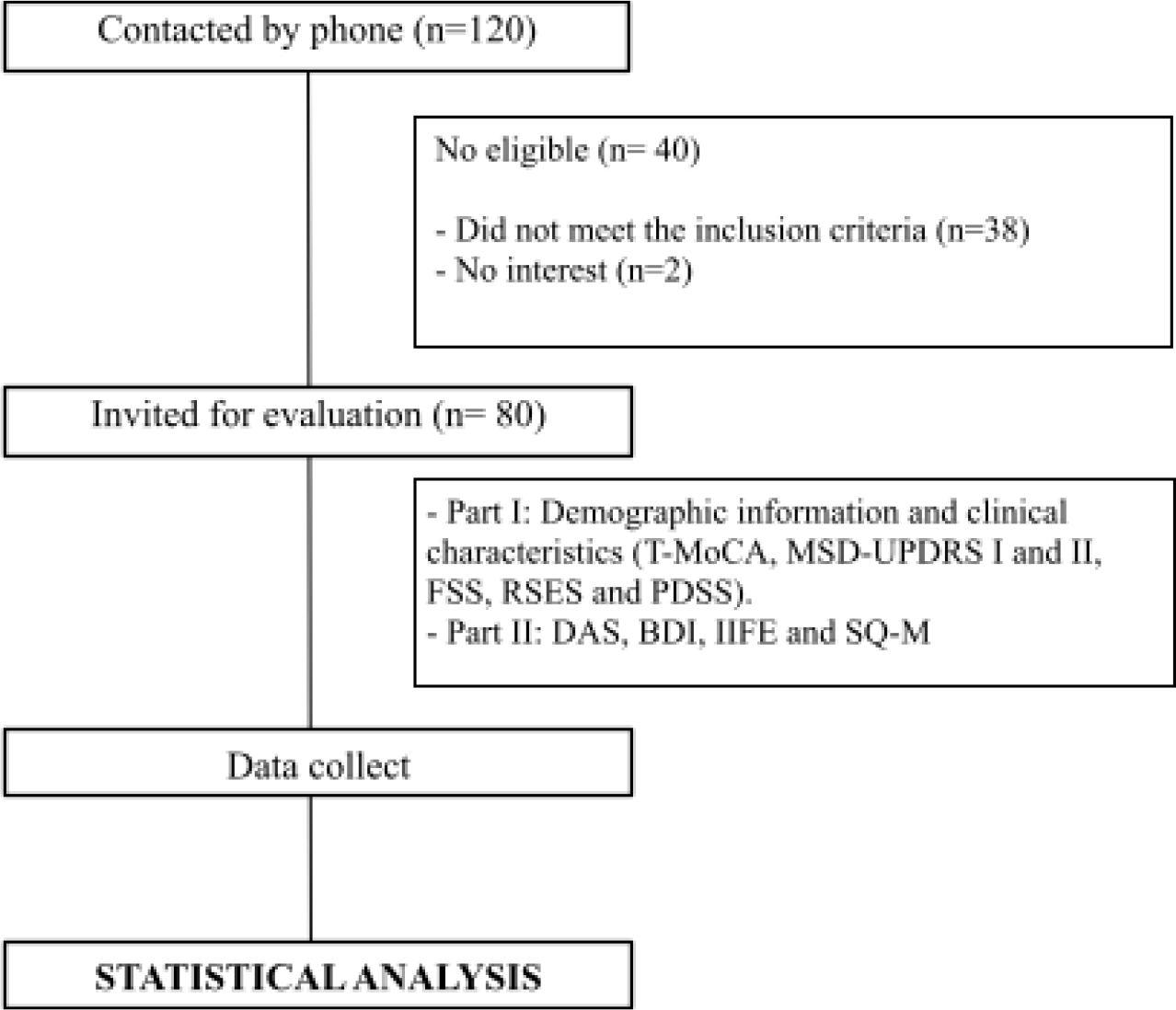

### Instruments

#### Telephone - Montreal Cognitive Assessment (T-MoCA)

The T-MoCA is an adapted version of the MoCA 30 administered by phone. It contains only the items that do not require the use of pencil and paper or visual stimuli, so its maximum score is 22 [41].

#### Movement Disorder Society - Unified Parkinson’s Disease Rating Scale (MDS-UPDRS)

The MDS-UPDRS is a tool to measure PD severity and progression based on the difficulties presented in the last seven days. In this study, only Part I and Part II were used [42].

#### Fatigue Severity Scale (FSS)

The FSS is a non-specific rating scale often used in PD. This instrument contains 9 questions with answers ranging from 1 (strongly disagree) to 7 (strongly agree). The total score is the mean of the 9 questions, and higher scores indicate higher fatigue degrees [43].

#### Rosenberg’s Self-esteem Scale (RSES)

The RSES comprises 10 questions to assess self-esteem. Questions are answered on a Likert-type scale of how many points range from strongly agree, agree, disagree, and strongly disagree [44].

#### Parkinson’s Disease Sleep Scale (PDSS)

The PDSS is a specific scale for assessing sleep disorders in PD [45]. It comprises 15 questions associated with sleep disorders based on the last week. The score ranges from always (0) to never (10), except for question 1, whose scale ranges from poor (0) to excellent (10). The maximum score is 150 [46].

#### Dyadic Adjustment Scale (DAS)

The DAS is a self-report tool to assess marital adjustment. It consists of 4 subscales of dyadic satisfaction, cohesion, consensus, and affectional expression, with scores ranging from 0 to 5. The total score (sum of all questions) ranges from 0 to 151. Higher scores indicate a better relationship. A total score below 101 indicates dissatisfaction [47].

#### Beck Depression Inventory (BDI)

The BDI is widely used to screen for depression and to measure behavioral manifestations and severity of depression. The BDI comprises 21 questions about quality and depressive symptoms, ranging from 0-63 points (0-13 no depression, 14-19 mild depression, 20-28 moderate, 29-63 severe depression). The total score is the sum of all values. The BDI has been described as a valid instrument to assess depressive symptoms in PD [48].

#### International Index of Erectile Function (IIEF) - Short-term SH

The 15-question International Index of Erectile Function (IIEF) Questionnaire is a validated, multidimensional, self-administered investigation. It has been recommended as a primary endpoint for clinical trials of ED and diagnostic evaluation of ED severity [49]. The questionnaire consists of 15 questions, grouped into five domains: erectile function, orgasm, sexual desire, sexual satisfaction, and general satisfaction. Each question has a value ranging from 1 to 5, and the sum of the answers generates a final score for each domain, with low values indicating poor quality of sexual life based on the last four weeks [50].

The ranges of scores allow classifying erectile dysfunction (ED) into five categories, based on the domain of erectile function, with scores ranging from 6 to 30: without ED (26-30); mild ED (22-25); mild to moderate ED (17-21); moderate ED (11-16); severe ED (6-10).

#### Sexual Quotient - Male (SQ-M) - Long-term SH

It was constructed and validated for the Brazilian population according to the sexual specificities of men. This quotient has 10 questions with five possible Likert-type answers: 0 (never); 1 (rarely); 2 (sometimes); 3 (approximately 50% of the time); 4 (most of the time); and 5 (always). The final score reflects the sexual performance pattern based on the last six months, which can be classified as having sexual dysfunction (<60 points) and without sexual dysfunction (≥60 points). This score is obtained by adding the numbers corresponding to each question and then multiplying by 2 [51].

### Statistical Analyses

Descriptive statistical analysis was used for demographic and clinical data. The Spearman Rank Order Correlation was used to test correlations among age, socio-economic classification, DDL, disease duration, disease onset, H&Y, T-MoCA, MDS-UPDRS I, MDS-UPDRS II, FSS, RSES, PDSS, DAS, BDI with IIEF total scores, EF domain, and SQ-M, and between EF domain of the IIEF and SQ-M.

A multiple regression model as predictor variables also included all factors that reached a significant statistical correlation with total scores of IIEF, EF domain, and SQ-M (response variable).

Additionally, the scores in IIEF and SQ-M were compared by Kolmogorov-Smirnov Test between participants with early onset PD (EOPD), i.e., diagnosed when aged less than 50 years and late-onset PD (LOPD), i.e., diagnosed when aged more than 50 years [52].

Differences were considered statistically significant for *p* < 0.05. The statistical analyses were performed using Statistica Version 13 (TIBCO Software Inc. USA).

## Results

The participants’ demographic and clinical characteristics are shown in Table 1.

**Table 1:** Demographic and clinical features of the participants.

| Variables | Mena | Std. Dev. |
| --- | --- | --- |
| Age | 53.55 | 10.82 |
| Socio-economic condition | 33.26 | 10.89 |
| Disease duration | 6.62 | 4.67 |
| H&Y | 1.86 | 0.68 |
| DDL | 566.87 | 429.52 |
| T-MoCA | 19.08 | 1.82 |
| MDS-UPDRS I | 12.81 | 8.40 |
| MDS-UPDRS II | 13.81 | 9.30 |
| FSS | 36.97 | 16.45 |
| PDSS | 98.31 | 25.28 |
| DAS | 110.03 | 9.44 |
| BDI | 11.93 | 8.90 |
| RSES | 16.45 | 16.45 |

H&Y stage: Hoehn and Yahr stage; DDL: Levodopa daily dose; T-MoCA: Telephone - Montreal Cognitive Assessment; MDS-UPDRS: Movement Disorder Society – Unified Parkinson's Disease Rating Scale; FSS: Fatigue Severity Scale; PDSS: Parkinson's Disease Sleep Scale; DAS: Dyadic Adjustment Scale; BDI: Beck Depression Inventory; RSES: Rosenberg's Self-esteem Scale.

Out of all the participants, it was found that 31% were in Stage 1, 51% were in Stage 2, and 18% were in Stage 3, according to the H&Y classification. Additionally, 68% of participants were less than 60 years old, and 64% presented EOPD. It is worth noting that none of the participants reported hypersexuality according to the last question of MDS-UPDRS I.

### Short-term sexual health (STSH)

The total score in IIEF, which investigated SH in the last four weeks, was 55.98±17.8. The results for each IIFE domain can be observed in Table 2.

**Table 2:**
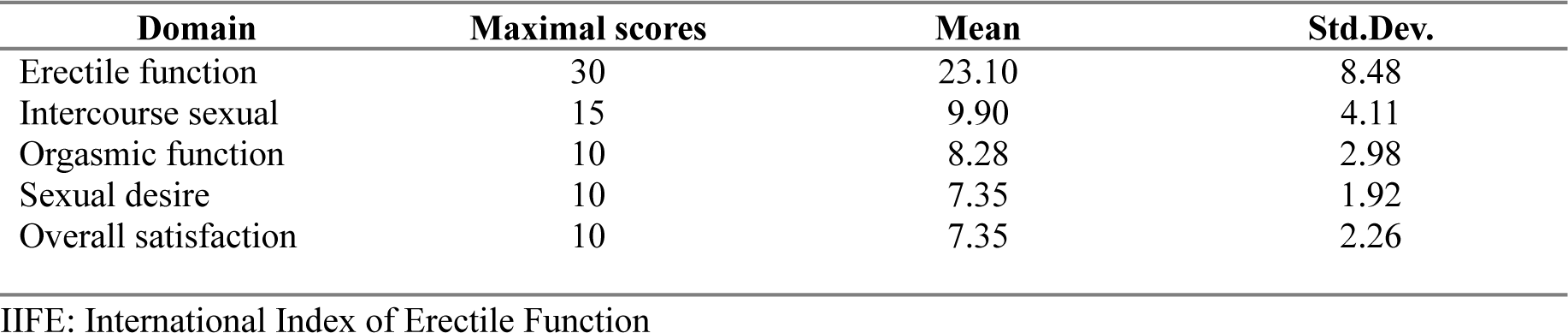
IIFE domain scores.

The total scores obtained by the IIEF had a weak positive statistically significant correlation with DAS score only (R=.28, p<.02). The regression model showed that DAS could predict IIEF with R^2^=.07 and Beta =.30.

The mean score in the erectile function (EF) domain was 23.10±8.48, below the minimal score adopted to identify erectile dysfunction (cut-off score <26).

The scores obtained by the EF domain had a moderate positive statistically significant correlation with T-MoCA and PDSS scores, a moderate negative statistically significant correlation with H&Y stages, MDS-UPDRS I and II, FSS, PDSS, and a weak negative statistically significant correlation with disease duration, RSES and BDI scores (Table 3).

**Table 3:** Correlation between EF, demographic and clinical variables.

| Variables | R | p |
| --- | --- | --- |
| Age | $R = -.202$ | $p = .071$ |
| Socio-economic condition | $R = .143$ | $p = .204$ |
| Disease duration | **$R = -.262$** | **$p = .018$** |
| H&Y | **$R = -.340$** | **$p = .001$** |
| DDL | $R = -.118$ | $p = .293$ |
| TMoCA | **$R = .359$** | **$p = .001$** |
| MDS-UPDRS I | $R = -.372$ | **$p = .000$** |
| MDS-UPDRS II | **$R = -.478$** | **$p = .000$** |
| FSS | **$R = -.320$** | **$p = .003$** |
| PDSS | **$R = .379$** | **$p = .000$** |
| DAS | $R = .115$ | $p = .362$ |
| BDI | **$R = -.233$** | **$p = .037$** |
| RSES | **R = -.268** | **p =.016** |

EF: Erectile function. H&Y stage: Hoehn and Yahr stage; DDL: Levodopa daily dose; T-MoCA: Telephone - Montreal Cognitive Assessment; MDS-UPDRS: Movement Disorder Society – Unified Parkinson's Disease Rating Scale; FSS: Fatigue Severity Scale; PDSS: Parkinson's Disease Sleep Scale; DAS: Dyadic Adjustment Scale; BDI: Beck Depression Inventory; RSES: Rosenberg's Self-esteem Scale.

The variables disease duration, T-MoCA, PDSS, H&Y stages, MDS-UPDRS I, and II, FSS, PDSS, RSES, and BDI were included in the multiple regression model, with only the MDS-UPDRS II remaining as a predictor variable for the EF, which presented R^2^=.31 and Beta =-.57.

### Long-term sexual health (LTSH)

The mean score in SQ-M, which investigated SH in the last six months, was 78.02±19.2, above the score adopted to identify sexual dysfunction (cut-off score<60). The mean scores by domain can be seen in Table 4.

**Table 4:** Correlation between SQ-M, demographic and clinical variables.

| Variables | R | p |
| --- | --- | --- |
| Age | R = -.117 | p =.299 |
| Socio-economic condition | R =.198 | p =.077 |
| Disease duration | R = -.154 | p =.171 |
| H&Y | **R= -.295** | **p=.007** |
| DDL | R = -.097 | p =.388 |
| T-MoCA | **R = .252** | **p =.024** |
| MDS-UPDRS I | **R = -.328** | **p =.002** |
| MDS-UPDRS II | **R = -.411** | **p =.000** |
| FSS | **R = -.343** | **p =.001** |
| PDSS | **R =.333** | **p =.002** |
| DAS | **R =.283** | **p =.023** |
| BDI | **R = -.290** | **p =.008** |
| RSES | **R = -.297** | **p =.007** |

SQ-M: Sexual Quotient – Male; H&Y stage: Hoehn and Yahr Stage; DDL: Levodopa daily dose; T-MoCA: Telephone - Montreal Cognitive Assessment; MDS-UPDRS: Movement Disorder Society – Unified Parkinson's Disease Rating Scale; FSS: Fatigue Severity Scale; PDSS: Parkinson's Disease Sleep Scale; DAS: Dyadic Adjustment Scale; BDI: Beck Depression Inventory. RSES: Rosenberg's Self-esteem Scale.

The total score obtained by SQ-M had a moderate positive statistically significant correlation with T-MoCA and DAS scores, a moderate positive statistically significant correlation with PDSS, a moderate negative statistically significant correlation with MDS-UPDRS I and II, FSS, PDSS, and a weak negative statistically significant correlation with H&Y stages, RSES and BDI scores (Table 4).

Furthermore, the multiple regression model included the variables T-MoCA, DAS, PDSS, MDS-UPDRS I, and II, FSS, PDSS, H&Y stages, RSES, BDI scores included in the multiple regression model, with only the BDI remaining in the final model as independent predictors for SQ-M with R^2^=.14, and Beta= -.40.

There was no significant difference between participants with EOPD and LOPD. Finally, a strong positive correlation was found between ED and SQ-M (r=.70; p<.000001).

## Discussion

SH is essential for maintaining overall well-being and health. Decreased SH has consequences that extend beyond the individual and can affect families and society [53]. However, there has been a lack of research on SH in people with PD. To the best of our knowledge, the present study was the first aiming to investigate the impact of the motor, non-motor, and social aspects on the short and long-term SH in MwPD by different but complementary instruments developed to assess SH.

Our results showed that motor, mental and social factors have a significant role in determining SH. Specifically, we found that the quality of the couple’s relationship had an impact on general short-term SH, motor disability level in daily life activities (DLA) had a negative impact on short-term SH, specifically on EF, while depression severity had a negative impact on long-term SH. Interestingly, we did not find any significant impact on SH from factors such as disease progression as measured by H&Y stage, age, disease onset, disease duration, and Levodopa daily dose (DDL), as well as non-motor symptoms severity. These findings suggest that addressing motor disability, mental health, and marital relationships may be crucial in improving the overall health of MwPD.

Taken together, several considerations can be made based on the results.

This study used two different instruments to assess SH in MwPD. In our study, we utilized two different instruments to evaluate SH in MwPD. The first tool, IIEF, assessed overall SH and specifically evaluated EF over the past four weeks. The second tool, SQ-M, took a qualitative approach and evaluated SH over the past six months. The EF domain of SQ-M was strongly correlated with each other and had a weak/moderate correlation with disease progression (H&Y stage), global cognitive capacity (T-MoCA), depression (BDI), self-esteem (RESS), motor and non-motor disability (MDS-UPDRS I and II), fatigue (FSS) and sleep (PDSS). However, only the IIFE total scores and SQ-M were correlated with the quality of couple relationships (DAS). Our multiple regression models revealed that different factors predicted scores on each instrument, with quality of marital relationship predicting general short-term SH (IIFE total score), motor disability predicting EF domain, and depression predicting SQ-M. Although the majority previous studies have used IIEF, primarily EF domain, [11,14,20,36,39] to evaluate SF in MwPD, our findings suggest that incorporating the SQ-M questionnaire can provide additional insight into how the PD affects SH.

Based on our results, many MwPD experience ED. Specifically, our findings show that 47% of MwPD reported ED based on IIEF, with 17% experiencing moderate to severe ED. Although age did not predict SH in the present study, it is important to note that 28% of participants under the age of 60 reported ED. Of these, 13% reported moderate to severe dysfunction. These findings align with previous studies that have identified ED as the most prevalent SD among MwPD, with prevalence rates ranging from 42.6% to 79%. [6,14,26,32,36]. Our results have demonstrated the importance of recognizing ED as a non-motor symptom in MwPD. We found that while several factors were associated with EF, it was only the motor disability in DLA that was able to predict the presence of ED. Previous research has also shown that individuals with PD who have more severe motor symptoms and disability tend to experience more SD [8,11,12,37,39], and ED [18], which is consistent with our findings. Several aspects evaluated by MDS-UPDRS part II can affect sexual performance as bed mobility, dressing ability, and hand coordination [6]. However, it is not entirely clear how motor disability affects erectile function. Our findings emphasize the need for clinicians to consider the impact of motor disability on sexual health in individuals with PD and highlight the demand for more studies to understand the relationship between motor disability and EF.

Our findings showed that the severity of depression plays a critical role in predicting long-term SH. This is consistent with previous research emphasizing the close link between depression and sexual dysfunction. In fact, studies conducted by Kotková and Weiss (2013), Vela-Desojo et al. (2020), Raciti et al. (2020), Özcan et al. (2015), Jitkritsadakul; Jagota, Bhidayasiri (2015) have also reported similar results. Additionally, our study identified depression severity as a significant predictor of SD in MwPD, which is consistent with the findings of Ferrucci et al. (2016) and Kotková and Weiss (2013) and Kummer, Cardoso, Teixeira (2009) that showed depression as a predictor for SD in men and women with PD. Specifically, for MwPD, higher levels of depression symptoms were associated with decreased sexual activity, as reported by Picillo et al. (2019). Since depression is a common non-motor symptom that may even be present before a diagnosis is possible, interventions to prevent or reduce depression are crucial to avoid or minimize the decreased SH in MwPD.

Although there was no specific cut-off for diagnosing SD based on the total score of IIEF, the mean score of 55 obtained was less than 75% of the maximum score of 75. Interestingly, it was found that the total score of IIEF was correlated and predicted by the quality of marital relationships only. This suggests that general aspects of short-term SH, extrapolating ED, are influenced by the complex aspects of marital relationship assessment by DAS. Previous studies have shown that the quality of the marital relationship is closely related to SH in both directions [8,34,54]. Our findings reinforce the importance of providing care not only for the patient but also for their partner.

Some previous studies have found associations between other non-motor symptoms and SD [55]. However, the MDS-UPDRS I, PDSS, and FSS, which assess general non-motor symptoms, sleep quality, and fatigue, respectively, although correlated with IIEF and SQ-M scores, had no predictive value for both in the present study. The same was observed for self-esteem: despite some features like drooling, excessive sweating, seborrhea, and hypomimia may decrease self-esteem [15], the RSES scores, although correlated with IIEF and SQ-M scores, had no predictive value for both.

Finally, although dopaminergic medication has been correlated with SD in MwPD [15], the DDL was not associated with SH in the present study.

The present study presents some strengths: using multidimensional generic instruments to assess SH, quality of couple relationships, depression, and cognition—Moreover, the use of disease-specific instruments that comprehensively assess motor and non-motor dysfunction in PD. The results obtained are promising and can serve as a foundation for further research in the field.

Our study has some limitations to be considered. One of the main limitations is that it was designed as cross-sectional, meaning that the information gathered was only based on data from a specific point in time. However, we tried to minimize this limitation by using different tools to assess the STSH and LTSH. Another limitation is that the sample size may not allow for large generalization to the PD population. However, we included participants from 5 different geographical areas with sizeable sociocultural differences, which may have helped to minimize this limitation. Lastly, we did not find men in advanced stages of PD who reported active sexual life to be included in the present study, which could also be considered a study limitation.

## Conclusion

SH was impaired in most MwPD, regardless of age, disease stage and disease onset. Quality of marital relationship predicts short-term SH, motor disability in DLA predicts ED, while depression severity predicts long-term SH. Therefore, an interdisciplinary approach to improve the quality of marital relationships, motor performance in DLA, and depressive mood should be available for MwPD since the early disease stages to avoid the decline in SH.

## Data Availability

All data produced in the present study are available upon reasonable request to the authors

## Glossary

SD: Sexual dysfunction
PWPD: People with Parkinson’s disease
SH: Sexual health
MwPD: Men with Parkinson’s disease
H&Y: Hoehn and Yahr
T-MoCA: Telephone – Montreal Cognitive Assessment
MDS-UPDRS: Movement Disorder Society – Unified Parkinson’s Rating Scale
FSS: Fatigue Severity Scale
RSES: Rosenberg’s Self-esteem Scale
PDSS: Parkinson’s Disease Sleep Scale
DAS: Dyadic Adjustment Scale
BDI: Beck Depression Inventory
IIFE: International Index of Erectile Function
SQ-M: Sexual Quotient-Male

## Acknowledgments

This article was produced as part of the activities of the FAPESP Research, Innovation, and Dissemination Center for Neuromathematics (grant #2013/ 07699-0, S. Paulo Research Foundation).

## Notes

### Competing Interest Statement

The authors have declared no competing interest.

### Funding Statement

This article was produced as part of the activities of the Center for Research, Innovation and Dissemination in Neuromathematics FAPESP (Process number 2013/07699-0, Sao Paulo State Research Support Foundation).

### Author Declarations

This study was approved by the Research Ethics Committee of the Federal University of Amapa (CAAE 39971420.0.0000.0003) and conducted in accordance with the Declaration of Helsinki.

